# Association of polypharmacy and burden of comorbidities on COVID-19 adverse outcomes in people with type 1 or type 2 diabetes

**DOI:** 10.1101/2023.08.12.23294016

**Authors:** JK Gupta, R Ravindrarajah, G Tilston, W Ollier, DM Ashcroft, AH Heald

## Abstract

**Aim:** To investigate whether polypharmacy and comorbidities conveyed more risk of adverse health outcomes following COVID-19 infection in people with type 1 diabetes (T1DM) or type 2 diabetes (T2DM).

**Materials and methods:** The Greater Manchester Care Record (GMCR) is an integrated database of electronic health records containing data collected from 433 general practices in Greater Manchester. Baseline demographic information (age, BMI, gender, ethnicity, smoking status, deprivation index), hospital admission or death within 28 days of infection were extracted for adults (18+) diagnosed with either T1DM or T2DM.

**Results:** For T2DM, 16 to 20 medications (p=0.005; OR [95% CI]=2.375 [1.306 to 4.319]) and > 20 medications (p<0.001; OR [95% CI]=3.141 [1.755 to 5.621]) were associated with increased risk of death following COVID-19 infection. Increased risk of hospital admissions in T2DM individuals was associated with 11 to 15 medications (p=0.013; OR [95% CI]=1.341 [1.063 to 1.692]) and above. This was independent of comorbidities, metabolic and demographic factors. For T1DM there was no association of polypharmacy with hospital admission. Respiratory, cardiovascular/cerebrovascular and gastrointestinal conditions were associated with increased risk of hospital admissions and deaths in T2DM (p<0.001).

**Conclusion:** We have shown in T2DM an independent association of multiple medications taken from 11 upwards with adverse health consequences following COVID-19 infection. We also found that individuals with diabetes develop comorbidities that were common across both T1DM and T2DM. This study has laid the foundation for future investigations into the way that complex pharmacological interactions may influence clinical outcomes in people with T2DM.

## Introduction

Acute Respiratory Syndrome coronavirus 2 (SARS-CoV-2) or COVID-19 is the pathogenic coronavirus that led to the 2020 pandemic. COVID-19 infections can lead to adverse outcomes, such as hospital admission or death, the risk of which are increased further for individuals diagnosed with either type 1 diabetes mellitus (T1DM) or type 2 diabetes mellitus (T2DM) ^1,2^. Increased risk of adverse health outcomes following COVID-19 infection have also been linked to other underlying medical conditions, which has raised concerns for chronic disease care ^3–7^. It has been showed that socially marginalised and psychiatrically vulnerable individuals are at higher risk of severe health outcomes following COVID-19 infection ^8^. McQueenie et al. (2020) investigated the UK Biobank data to determine the association between multi-morbidity (including polypharmacy as a proxy) and COVID-19 infection risk ^9^. McQueenie et al. (2020) reported that individuals diagnosed with ≥2 cardiometabolic conditions and increasing polypharmacy were associated with increased the risk of COVID-19 infection ^9^.

Previous studies of COVID-19 in individuals with T1DM and T2DM (in the Greater Manchester (GM) area) have shown that the prescribing of certain medications influenced the likelihood of hospitalisation or death following COVID-19 infection ^10,11^. Many individuals in this population are often prescribed, and are taking, multiple medications as they live with other long-term conditions. The definition of ‘polypharmacy’ has been shown to widely vary in the published literature ^12^. In addition to this term, Masnoon et al. (2017) summarised that the terms minor, moderate and major polypharmacy were used in literature to describe when between 2 to 11 or more medications are consumed. The most commonly reported number of medications across these ‘polypharmacy’ terms was greater than or equal to 5, as identified in the review ^12^.

It is widely accepted that the higher the number of medications prescribed and taken by an individual, the higher the risk of poor health outcomes. Recent reviews have reported a high prevalence of polypharmacy in older people diagnosed with diabetes and an association with several health-related outcomes, including falls, syncope, hospitalization, and death, as well as highlighting the need to reduce inappropriate prescribing ^13–15^. Several studies have also investigated the relationship between polypharmacy and severe COVID-19 health outcomes ^16,17^. However, there is little understanding of how polypharmacy in people with diabetes might affect their risk of severe health outcome post COVID-19 infection, other than the public health burden and impact of adverse drug-drug interactions in individuals ^18^.

Electronic health records (EHR) emerged as a useful tool for public health and COVID-19 research, as described by Madhavan et al. (2021) and Casey et al. (2016) ^19,20^. It is also useful for improving health service for example by identifying and understanding health inequalities ^21,22^.

To date, there are no studies that explore the associations between both polypharmacy and comorbidity with adverse health outcomes, such as hospital admission or death, post first infection of COVID-19 specifically in individuals with diabetes. This retrospective cohort study aimed to investigate whether polypharmacy independent of comorbidity conveys greater risk of adverse outcomes in people with diabetes when testing positive for COVID-19 infection in UK EHR data.

## Materials and methods

### Cohort data source

The Greater Manchester Care Record (GMCR) is an integrated database of primary care, secondary care and mental health trusts from across GM (https://gmwearebettertogether.com/research-and-planning/) for analyses covering a population of approximately 3 million people. Health and care data were collected from 433 of 435 (99.5%) general practices in GM. Data were de-identified at source and were extracted from the GMCR database. This was a retrospective cohort study with the period of follow-up 2020 to 2022 in relation to the main impact period of the COVID-19 pandemic. The inclusion criteria for this study were defined as individuals that are registered with a GM GP practice and with a diagnosis of T1DM or T2DM and age 18 or above. Individuals with a positive test for COVID-19 close to January 2020 were included in this study. The study included all people diagnosed with diabetes and with data available in the GMCR in January 2020. No power calculation was performed as we included all people with that diagnosis.

### Variables and data cleaning

Baseline demographic information (age, BMI, gender, ethnicity, smoking status, deprivation index), hospital admission or death within 28 days of infection were extracted for adults (18+) diagnosed with either T1DM or T2DM (the codes applied are summarised in Appendix 1). Other rare forms of diabetes were not included.

Hospital admissions were recorded within 4 weeks after, or 2 weeks before a positive COVID-19 test (between Jan 2020 to May 2022). The exposure was defined as prescribed medications, which were recorded in the EHRs and mapped to the corresponding BNF (British National Formulary) chapters. For this study, medications were considered at a single point in time, the month closest to first COVID-19 infection date (January 2020 onwards). The BNF groups were not mutually exclusive, therefore medications that are grouped under more than one chapter were counted. History of comorbidity was collected before March 2020.

Individuals who were not assigned a gender were excluded. A total of 410 individuals having a code for both T1DM and T2DM were excluded. The study confounders were determined by literature review. In studies using real world data analysis, a number of factors were not covered in the coded data such as household makeup and employment. Links to the codes used for diabetes can be found in the supplementary information. Data were checked for extreme outliers or inconsistent values and removed as appropriate.

### Ethics

This project was reviewed, and ethical approval for COVID-19 research was overseen by Health Innovation Manchester and granted by the Greater Manchester Care Record (GMCR) review board (ref: IDCR-RQ-046). This research was performed with anonymised data, in line with the Health Research Authority’s Governance arrangements for research ethics committees.

### Statistical methods

Multivariable logistic regression analyses using a forward stepwise approach were performed on the T1DM and T2DM individuals, measuring exposure to number of medications and comorbidities, with either hospital admission or death after COVID-19 infection (within 28 days of COVID-19 diagnosis) as the outcome. The models were adjusted for age, BMI, ethnicity, smoking status, IMD, eGFR <60 ml/min/1.73m^2^, cholesterol, HBA1C and blood pressure. Analyses were performed in STATA v17. This manuscript follows the reporting recommendation of RECORD-PE ^23^.

## Results

The study cohort included patients diagnosed as T1DM and T2DM separately. Across the Greater Manchester Region a total of 145,907 individuals were diagnosed with T2DM and 9,705 were diagnosed with T1DM (Table 1; Table 2). The number of deaths in T1DM were too low (n=30) for further statistical analyses to explore the association with mortality.

**Table 1.** Baseline demographics of individual diagnosed with Type 1 diabetes (T1DM) or Type 2 diabetes (T2DM).

|  | Type 1 Diabetes<br>*N*=9,705 | Type 2 Diabetes<br>*N*=145,907 |
| --- | --- | --- |
| Age (years) | 47.3±16.8 | 65.8±13.8 |
| BMI (kg/m<sup>2</sup>) | 27.7±7.0 | 31.0±7.7 |
| **Sex** |  |  |
| Female | 4143 (42.7) | 65922 (45.2) |
| Male | 5562 (57.3) | 79985 (54.8) |
| **Smoking status** |  |  |
| Non-smoker | 3967 (40.9) | 51823 (35.5) |
| Current smoker | 1804 (18.6) | 20906 (14.3) |
| Ex-smoker | 3765 (38.8) | 71519 (49.0) |
| Unknown/missing | 169 (1.7) | 1659 (1.1) |
| **Ethnicity** |  |  |
| White | 8020 (82.6) | 102051 (69.9) |
| Asian | 565 (5.8) | 25359 (17.4) |
| Black | 221 (2.3) | 5176 (3.6) |
| Mixed | 107 (1.1) | 1528 (1.1) |
| Other | 379 (3.9) | 6184 (4.2) |
| Missing/refused | 413 (4.3) | 5609 (3.8) |
| **Deprivation index/Townsend quintile (IMD)** |  |  |
| 1 (least deprived) | 1182 (12.2) | 13471 (9.2) |
| 2 | 1451 (15.0) | 19080 (13.1) |
| 3 | 1395 (14.4) | 18017 (12.4) |
| 4 | 1984 (20.4) | 28863 (19.8) |
| 5 (most deprived) | 3680 (37.9) | 66426 (45.5) |
| Missing | 13 (0.1) | 50 (0.03) |
| **Total number of medications prescribed \*** |  |  |
| 0 | 543 (5.60) | 13359 (9.16) |
| 1 to 5 | 5822 (59.99) | 51999 (35.64) |
| 6 to 10 | 1879 (19.36) | 38533 (26.41) |
| 11 to 15 | 744 (7.67) | 21433 (14.69) |
| 16 to 20 | 348 (3.59) | 9880 (6.77) |
| > 20 | 369 (3.80) | 10703 (7.34) |
| Systolic blood pressure (mm Hg) † | 127.1±15.2 | 131.1±14.5 |
| Diastolic blood pressure (mm Hg) † | 74.8±9.8 | 75.9±9.6 |
| HBA1C (mmol/mol) † | 69.3±20.3 | 56.8±17.0 |
| eGFR (mL/min/1.73 m<sup>2</sup>) < 60 ‡ | 907 (9.4) | 25851 (17.7) |
| eGFR (mL/min/1.73 m<sup>2</sup>) † | 81.0±15.5 | 75.4±17.2 |
| Cholesterol (mmol/mol) † | 4.5±1.1 | 4.3±1.1 |
| LDL (mmol/L) † | 2.4±0.9 | 2.2±0.9 |
| HDL (mmol/L) † | 1.5±0.5 | 1.2±0.4 |
| Diabetes duration (years) | 20.6±14.3 | 10.4±7.45 |
| **Vaccination status: (defined prior to/around the time of first infection)** |  |  |
| Yes | 8816 (90.8) | 135141 (92.6) |
| No | 889 (9.2) | 10766 (7.4) |
| **Prescribed medications: \*** |  |  |
| Metformin | 1015 (10.5) | 72084 (49.4) |
| SGLT2 • | 163 (1.7) | 16134 (11.1) |
| Insulin | 6854 (70.6) | 12149 (8.3) |
| Arb/ACE inhibitors | 1759 (18.1) | 44127 (30.2) |
| Antiplatelet (clopidogrel/aspirin) | 1177 (12.1) | 31403 (21.5) |
| Angiotensin receptor blockers | 710 (7.3) | 20168 (13.8) |
| Sulphonylureas | 98 (1.0) | 22011 (15.1) |
| GLP1 receptor agonists | 79 (0.8) | 4475 (3.1) |
| Alogliptin | 36 (0.4) | 10196 (7.0) |
| Linagliptin | 37 (0.4) | 5388 (3.7) |
| Saxagliptin | < 10 (0) | 944 (0.7) |
| Sitagliptin | 29 (0.3) | 6570 (4.5) |
| Vildagliptin | < 10 (0) | 396 (0.27) |
| **Admission to hospital (within 28 days of infection)** | 146(1.5) | 2107 (1.4) |
| Length of stay in hospital (days) | 4.5±8.3 | 7.1±11.9 |
| **Deaths (within 28 days of confirmed infection)** | 30 (0.3) | 885 (0.6) |

Categorical variables presented as N (%) and continuous variables as mean (±SD)
Townsend index: 1 denotes the least deprived and 5 the most deprived.
\* Medications prescribed close to January 2020.
† Closest point before first COVID-19 +ve test
‡ Estimated glomerular filtration rate
• Sodium glucose co-transporter 2 inhibitor (SGLT2i) or a glucagon-like peptide 1 (GLP-1) agonist.

**Table 2.** Prevalence of comorbidity groups in individuals diagnosed with T2DM. The top 6 groups are shown in italics.

| Morbidity group | N | % |
| --- | --- | --- |
| *Hypertension* | 101995 | 69.9 |
| *GI/liver disease* | 82193 | 56.3 |
| *Mental health disorder* | 65487 | 44.9 |
| *Pain* | 60237 | 41.3 |
| *Cardiovascular/cerebrovascular* | 48213 | 33.0 |
| *Respiratory/sinus* | 41653 | 28.6 |
| Sensory | 34541 | 23.7 |
| Skin/connective tissue disorder | 28459 | 19.5 |
| Chronic kidney disease | 27931 | 19.1 |
| Neurological disorders | 22795 | 15.6 |
| Thyroid disorders | 14476 | 9.9 |
| Prostate diseases | 9276 | 6.4 |
| Alcohol substance abuse | 9108 | 6.2 |
| Cancer | 7892 | 5.4 |
| Dementia | 6498 | 4.5 |
| Glaucoma | 6262 | 4.3 |
| Learning disability | 1016 | 0.7 |

A varying number of multiple medications were prescribed to individuals with T1DM or T2DM (Table 1). A large proportion of individuals were prescribed between 1 to 5 medications; the distribution of number of medications prescribed and BNF chapters can be seen in Figure S1 and Figure S2. In T2DM, after endocrine and cardiovascular medications, the largest number of multiple prescriptions were from the gastrointestinal (GI) BNF chapter.

Common comorbidities were grouped into broader categories (see Supplementary Information Table S1) and counts were collated for T1DM and T2DM. The top six modal comorbidity groups identified in T2DM were mental health conditions, hypertension, gastrointestinal or liver disease, pain, respiratory conditions (or sinus-related) and cardiovascular/cerebrovascular conditions (Table 2). Of the individuals diagnosed with T2DM, 69.9% were diagnosed with hypertension and 44.9% with mental health conditions (Table 2). The same six comorbidity groups were also identified in the individuals with T1DM (Table S2).

### Logistic Regression Analysis

Odds ratios (95% confidence intervals) of the univariate regression are reported in the individuals with T1DM and T2DM respectively.

### T1DM

An increased risk of adverse outcomes post-infection was observed for individuals with T1DM and GI/liver disorders (p<0.001; OR [95% CI] = 3.452 [2.118 to 5.625]) and pain associated conditions (p=0.04; OR [95% CI] = 1.556 [1.021 to 2.372]) (Table 3). Age was determined as slightly protective of hospital admission following COVID-19 diagnosis (p<0.001; OR [95% CI] = 0.964 [0.949 to 0.980]). Other significant variables included eGFR < 60 (p<0.001; OR [95% CI] = 2.794 [1.746 to 4.47]). There was no association with polypharmacy.

**Table 3.** Logistic regression analyses of hospital admission following COVID-19 diagnosis in individuals with T1DM.

| Variable | OR (95% CI) | p value |
| --- | --- | --- |
| Age | 0.964 (0.949 to 0.980) | < 0.001 |
| Sex | 0.792 (0.537 to 1.167) | 0.239 |
| BMI | 0.989 (0.960 to 1.02) | 0.485 |
| **Ethnicity** |  |  |
| Asian | 0.919 (0.431 to 1.956) | 0.826 |
| Black | 0.388 (0.052 to 2.882) | 0.355 |
| Mixed | 1.889 (0.444 to 8.04) | 0.390 |
| Other | 0.447 (0.108 to 1.852) | 0.267 |
| Unknown | 0.250 (0.035 to 1.809) | 0.170 |
| **Smoking status** |  |  |
| Current smoker | 0.856 (0.487 to 1.507) | 0.590 |
| Ex- smoker | 1.022 (0.665 to 1.571) | 0.921 |
| Unknown/missing | 1.237 (0.285 to 5.359) | 0.777 |
| **IMD** |  |  |
| 2 | 0.865 (0.404 to 1.851) | 0.708 |
| 3 | 0.65 (0.291 to 1.452) | 0.294 |
| 4 | 0.999 (0.501 to 1.992) | 0.998 |
| 5 (most deprived) | 0.843 (0.441 to 1.61) | 0.604 |
| eGFR<60 | 2.794 (1.746 to 4.47) | < 0.001 |
| Cholesterol | 0.925 (0.780 to 1.097) | 0.369 |
| HBA1C | 1.002 (0.993 to 1.011) | 0.681 |
| Systolic BP | 0.998 (0.984 to 1.012) | 0.788 |
| Diastolic BP | 1.018 (0.996 to 1.041) | 0.105 |
| **Comorbidity groups** |  |  |
| Mental health disorder | 0.690 (0.463 to 1.028) | 0.068 |
| Hypertension | 1.179 (0.746 to 1.865) | 0.480 |
| GI/liver disorder | 3.452 (2.118 to 5.625) | < 0.001 |
| Pain | 1.556 (1.021 to 2.372) | 0.040 |
| Respiratory/sinus conditions | 0.986 (0.650 to 1.496) | 0.948 |
| Cardiovascular/cerebrovascular | 1.327 (0.814 to 2.162) | 0.257 |
| **Total number of medications** |  |  |
| 1 to 5 | 1.001 (0.235 to 4.271) | 0.999 |
| 6 to 10 | 2.139 (0.488 to 9.381) | 0.313 |
| 11 to 15 | 2.496 (0.539 to 11.554) | 0.242 |
| 16 to 20 | 3.577 (0.738 to 17.329) | 0.113 |
| > 20 | 2.158 (0.424 to 10.989) | 0.355 |

Odds ratios and 95% confidence intervals from the logistic regression model of hospital admission post COVID-19 diagnosis.

### T2DM

In individuals with T2DM, an increased number of medications, from 11 upwards was associated with increased risk of hospital admission following COVID-19 infection (Table 4). Individuals diagnosed with co-morbidities in the following morbidity groups had an increased likelihood of hospital admission: mental health disorders (p=0.003; OR [95% CI] = 1.167 [1.054 to 1.292]); Gastro-intestinal/liver disorders (p<0.001; OR [95% CI] = 1.977 [1.741 to 2.245]); pain (p<0.001; OR [95% CI] = 1.407 [1.265 to 1.564]); respiratory/sinus conditions (p<0.001; OR [95% CI] = 1.235 [1.115 to 1.369]) and cardiovascular/cerebrovascular conditions (p<0.001; OR [95% CI] = 1.443 [1.292 to 1.612]). People with T2DM and eGFR level <60 (p<0.001; OR [95% CI] = 1.431 [1.268 to 1.614]) also had an increased risk of hospital admission, similarly HBA1C levels (p=0.009; OR [95% CI] = 1.004 [1.001 to 1.006]) were also at increased risk (Table 4).

**Table 4.** Logistic regression analyses of hospital admission within 28 days of COVID-19 diagnosis in individuals with T2DM.

| Variable | OR (95% CI) | p value |
| --- | --- | --- |
| Age | 0.993 (0.988 to 0.998) | 0.009 |
| Sex | 1.312 (1.181 to 1.457) | < 0.001 |
| BMI | 1.011 (1.004 to 1.018) | 0.002 |
| **Ethnicity** |  |  |
| Asian | 1.213 (1.058 to 1.39) | 0.006 |
| Black | 1.441 (1.129 to 1.838) | 0.003 |
| Mixed | 1.059 (0.632 to 1.774) | 0.827 |
| Other | 0.96 (0.744 to 1.239) | 0.754 |
| Unknown | 0.947 (0.701 to 1.28) | 0.725 |
| **Smoking status** |  |  |
| Current smoker | 0.507 (0.419 to 0.614) | < 0.001 |
| Ex- smoker | 0.998 (0.894 to 1.115) | 0.977 |
| Unknown/missing | 0.856 (0.525 to 1.396) | 0.533 |
| **IMD** |  |  |
| 2 | 1.07 (0.851 to 1.345) | 0.562 |
| 3 | 1.091 (0.868 to 1.373) | 0.454 |
| 4 | 1.278 (1.039 to 1.572) | 0.020 |
| 5 (most deprived) | 1.236 (1.017 to 1.503) | 0.033 |
| eGFR<60 | 1.431 (1.268 to 1.614) | < 0.001 |
| Cholesterol | 1.021 (0.976 to 1.068) | 0.369 |
| HbA1C | 1.004 (1.001 to 1.006) | 0.009 |
| Systolic BP | 1.001 (0.997 to 1.005) | 0.709 |
| Diastolic BP | 1.002 (0.996 to 1.008) | 0.470 |
| **Comorbidity groups** |  |  |
| Mental health disorder | 1.167 (1.054 to 1.292) | 0.003 |
| Hypertension | 1.074 (0.946 to 1.219) | 0.272 |
| GI/liver disorder | 1.977 (1.741 to 2.245) | < 0.001 |
| Pain | 1.407 (1.265 to 1.564) | < 0.001 |
| Respiratory/sinus conditions | 1.235 (1.115 to 1.369) | < 0.001 |
| Cardiovascular/cerebrovascular | 1.443 (1.292 to 1.612) | < 0.001 |
| **Total number of medications** |  |  |
| 1 to 5 | 1.123 (0.901 to 1.401) | 0.302 |
| 6 to 10 | 1.108 (0.886 to 1.387) | 0.370 |
| 11 to 15 | 1.341 (1.063 to 1.692) | 0.013 |
| 16 to 20 | 1.45 (1.124 to 1.87) | 0.004 |
| > 20 | 1.53 (1.191 to 1.966) | 0.001 |

Odds ratios and 95% confidence intervals from the logistic regression model of hospital admission post COVID-19 diagnosis.

Males were more likely to be admitted to hospital after infection with COVID-19 (p<0.001; OR [95% CI] = 1.312 [1.181 to 1.457]). High BMI was associated with a higher likelihood of adverse outcomes (hospitalisation & mortality) from COVID-19 (p=0.002; OR [95% CI] = 1.011 [1.004 to 1.018]). Age was protective of hospital admission; however the odds ratio was only slightly less than 1 (p=0.009; OR [95% CI] = 0.993 [0.988 to 0.998]). Individuals of Asian (p=0.006; OR [95% CI] = 1.213 [1.058 to 1.39]) or Black (p=0.003; OR [95% CI] = 1.441 [1.129 to 1.838]) ethnicities were also more likely to be admitted to hospital following infection (Table 4).

Individuals who identified as ‘current smokers’ were not identified as being at increased risk (p<0.001; OR [95% CI] = 0.507 [0.419 to 0.614]). However, individuals who lived in areas with higher social disadvantage were more likely to be admitted to hospital - IMD quintiles 4 and 5 (p=0.02; OR [95% CI] = 1.278 [1.039 to 1.572]; p=0.033; OR [95% CI] = 1.236 [1.017 to 1.503]) (Table 4).

In individuals with T2DM, the number of deaths was 885 (0.6% of the total number of individuals diagnosed with T2DM) (Table 1). An increased number of medications, from 16 to 20 to > 20 was associated with increased risk of death following COVID-19 infection (p=0.005; OR [95% CI] = 2.375 [1.306 to 4.319] and p<0.001; OR [95% CI] = 3.141 [1.755 to 5.621] respectively) (Table 5).

**Table 5.** Logistic regression analyses of deaths within 28 days of COVID-19 diagnosis in individuals with T2DM.

| Variable | OR (95% CI) | p value |
| --- | --- | --- |
| Age | 1.092 (1.078 to 1.107) | < 0.001 |
| Sex | 1.35 (1.054 to 1.728) | 0.017 |
| BMI | 1.021 (1.007 to 1.036) | 0.004 |
| **Ethnicity** |  |  |
| Asian | 0.719 (0.46 to 1.123) | 0.147 |
| Black | 0.666 (0.272 to 1.631) | 0.374 |
| Mixed | 1.418 (0.447 to 4.5) | 0.553 |
| Other | 0.696 (0.357 to 1.36) | 0.289 |
| **Smoking status** |  |  |
| Unknown | 1.259 (0.717 to 2.212) | 0.423 |
| Current smoker | 1.015 (0.665 to 1.549) | 0.945 |
| Ex- smoker | 0.945 (0.724 to 1.234) | 0.677 |
| Unknown/missing | Omitted |  |
| **IMD** |  |  |
| 2 | 1.444 (0.88 to 2.37) | 0.146 |
| 3 | 1.181 (0.7 to 1.993) | 0.534 |
| 4 | 1.481 (0.921 to 2.381) | 0.105 |
| 5 (most deprived) | 1.684 (1.081 to 2.623) | 0.021 |
| eGFR<60 | 1.382 (1.078 to 1.771) | 0.011 |
| Cholesterol | 1.084 (0.973 to 1.207) | 0.145 |
| HBA1C | 1 (0.992 to 1.008) | 0.967 |
| Systolic BP | 0.993 (0.985 to 1.002) | 0.122 |
| Diastolic BP | 1.001 (0.987 to 1.015) | 0.906 |
| **Comorbidity groups** |  |  |
| Mental health disorder | 1.544 (1.219 to 1.954) | < 0.001 |
| Hypertension | 0.794 (0.574 to 1.099) | 0.164 |
| GI/liver disorder | 0.519 (0.404 to 0.666) | < 0.001 |
| Pain | 0.513 (0.4 to 0.657) | < 0.001 |
| Respiratory/sinus conditions | 1.573 (1.245 to 1.988) | < 0.001 |
| Cardiovascular/cerebrovascular | 1.691 (1.297 to 2.203) | < 0.001 |
| **Total number of medications** |  |  |
| 1 to 5 | 0.583 (0.32 to 1.06) | 0.077 |
| 6 to 10 | 0.945 (0.536 to 1.665) | 0.844 |
| 11 to 15 | 1.429 (0.802 to 2.545) | 0.226 |
| 16 to 20 | 2.375 (1.306 to 4.319) | 0.005 |
| > 20 | 3.141 (1.755 to 5.621) | < 0.001 |

Odds ratios and 95% confidence intervals from the logistic regression model of mortality post COVID-19 diagnosis.

People diagnosed with the following long-term conditions were at higher risk of death post-infection: mental health disorders (p<0.001; OR [95% CI] = 1.544 [1.219 to 1.954]); respiratory/sinus conditions (p<0.001; OR [95% CI] = 1.573 [1.245 to 1.988]) or cardiovascular/cerebrovascular conditions (p<0.001; OR [95% CI] = 1.691 [1.297 to 2.203]). A protective effect was observed for individuals living with GI/liver disorder (p<0.001; OR [95% CI] = 0.519 [0.404 to 0.666]) or pain (p<0.001; OR [95% CI] = 0.513 [0.4 to 0.657]) (Table 5).

As with hospital admission, the risk of death increased in males and with high BMI (p=0.017; OR [95% CI] = 1.35 [1.054 to 1.728] and p=0.004; OR [95% CI] = 1.021 [1.007 to 1.036] respectively). Age was associated with an increased risk of mortality post COVID-19 infection (p<0.001; OR [95% CI] = 1.092 [1.078 to 1.107]) (Table 5).

Individuals with T2DM and living in the most deprived regions (IMD 5) were at higher risk of death following COVID-19 infection (p=0.021; OR [95% CI] = 1.684 [1.081 to 2.623]) or if their eGFR level measure was <60 (p=0.011; OR [95% CI] = 1.382 [1.078 to 1.771]) (Table 5).

## Discussion

This study uniquely explored the relationship between number of medications and adverse outcome from COVID-19 infection in individuals diagnosed with diabetes in the Greater Manchester population and is therefore distinct from our previous published studies in this area ^10,11^. We found that the number of medications prescribed to an individual was associated with an increased risk of severe outcome from COVID-19 infection, particularly in people living with T2DM (Table 4; Table 5). Given the large proportion of individuals diagnosed with comorbidities, it was not unexpected that many of these individuals were prescribed multiple medications (Figure S1; Figure S2). However, the effect of multiple medications was independent of the presence of the most common comorbidities. Polypharmacy, independent of other factors including major multimorbidity, was associated with an increased likelihood of hospital admission in people with T1DM and of hospitalisation and death in T2DM following COVID-19 infection. This has not been specifically reported previously, although it was described in older individuals ^24^. This is of relevance to the consequences following any serious viral infection in people with diabetes, while also highlighting the importance of regular medicine reviews in everyone with diabetes, where there may be an opportunity to reduce the prescribed medications ^25^.

The mean ages of individuals diagnosed with T1DM or T2DM are similar to the mean ages of a Swedish cohort investigated by ^1^. In our study we also identified several co-morbidities that increased the risk of adverse health outcome post COVID-19 infection. Similar co-morbidities were identified for both individuals with T1DM and T2DM. Notably, we also identified that there were a large proportion of diagnoses of mental health conditions in people with T2DM (Table 2), though the highest number of diagnoses was observed in people with T1DM (Table S2). The relation between physical health conditions and poor mental health is an area that is not fully understood. In addition to this, the impact of polypharmacy is unknown.

The risk of hospital admission in current smokers was less than non-smokers. The reason for this may be that people with multiple health issues and who are smokers may have stopped smoking in the weeks or months before a COVID-19 infection and therefore not be deemed as a smoker at the time of the COVID-19 infection.

Over-prescribing is an issue in the current healthcare system. Medication is routinely recorded, therefore in clinical practice there is the potential to flag the number of medications prescribed to clinicians to monitor. Thereby the number of prescriptions given to vulnerable individuals could be limited as appropriate and effective for the individual, though this does require both routine monitoring and structured reviews of the medications that people are taking. The UK healthcare system looks to improve the management of prescriptions, and the removal of unnecessary medications is key to reducing the burden of polypharmacy ^26,27^.

One of the limitations of this study is that a measure of ‘frailty’ was not included ^28^. However, a count of comorbidities is included which could possibly be used as a proxy for frailty. As the data from this study originated from electronic health records, it is also subject to the limitations of how information is coded when originally entered into the systems, such as missing data. Only 70% of individuals with T1DM were recorded with a prescription of insulin (Table 1), this could be due to lack of recording or unclear diagnosis, which highlights another issue with routinely collected data. Another issue is that medications are recorded in the health care records are not true reflections of medications that are concordant with the choices of the individual ^29^. A study in the United States demonstrated that EHR-related medication errors can occur at different stages including at ordering, preparation, dispensing, administering or monitoring stage, these can ultimately affect the data in EHRs ^30^. Inaccuracies of coding are inherent in any project that relies on primary coded data. Nevertheless, the large number of people included in the study means that such inaccuracies are unlikely materially to influence the results. Furthermore, medications can also be counted in more than one BNF chapter, making it difficult to determine unique counts. As this data was from one region in the UK, it is difficult to generalise to the rest of the population. Finally in studies using real world data analysis, there will be a number of factors that are not covered in the coded data such as household makeup and employment.

Future work would include investigating the modal medications prescribed in this cohort and explore possible drug-drug interactions between medications that could contribute to increasing the risk of adverse outcome post COVID-19 infection. The burden of mental health conditions on people with diabetes and how to effectively manage multiple long-term conditions, could also be explored. The knowledge gained would be useful for future pandemics and better prepare our healthcare system. In conclusion, we have identified that multiple medications were prescribed to many people living with diabetes and that this was associated with a higher risk of adverse health outcome following COVID-19 infection. We also found that individuals with diabetes also developed comorbidities that were common across both Type 1 and 2 diabetes. Our analysis confirmed the impact of higher levels of deprivation on increasing a person’s a risk of adverse outcome post COVID-19 infection. This study has laid the foundation for future investigations into the increased and complex treatments people living with diabetes who develop other clinical complications.

## Data Availability

The datasets generated during and analysed during the current study are not publicly available as all data access is subject to review by Health Innovation Manchester.

## Acknowledgements

This work was supported by the Turing-Manchester Feasibility Project Funding. The authors recognise the Greater Manchester Care Record (a partnership of Greater Manchester Health and Social Care Partnership, Health Innovation Manchester and Graphnet Health, on behalf of Greater Manchester localities) in the provision of data required to undertake this work. This work uses data provided by patients and collected by the NHS as part of their care and support. GT was funded by the NIHR Manchester Biomedical Research Centre (NIHR203308) for this work. DMA is supported by the NIHR Greater Manchester Patient Safety Research Collaboration (PSRC). The views expressed are those of the authors and not necessarily those of the NIHR or the Department of Health and Social Care.

## Author contributions

RR performed the data cleaning and data analyses. JG, WO and AHH conceived the study. JG prepared the figures and the manuscript. GT extracted the original dataset. DM WO and AHH contributed to all sections of the paper and DM provided expert input from a pharmacological perspective. All authors were involved in designing the study, interpreting results and the reviewing and editing of the manuscript.

## Supplementary Materials

Links to the clinical codes applied for diabetes diagnoses can be found in the ‘methods’ section of the supplementary material file. Figure S1: distribution of medications prescribed in T1DM individuals. Figure S2: distribution of medications prescribed in T1DM individuals. Table S1: breakdown of comorbidity groups. Table S2: Prevalence of comorbidities groups in T1DM.

## Preprint

A preprint has previously been published [31].

