## Supplementary Information for "Association of polypharmacy and burden of comorbidities on COVID-19 adverse outcomes in people with type 1 or type 2 diabetes"

**Methods**

The following diabetes codes were applied: <https://github.com/rw251/gm-idcr/tree/master/shared/clinical-code-sets/conditions/diabetes-type-i/1> and <https://github.com/rw251/gm-idcr/tree/master/shared/clinical-code-sets/conditions/diabetes-type-ii/1>.

**Results**

Number of people with T1DM

| BNF chapters | 0 | 1 to 5 | 6 to 10 | 11 to 15 | 16 to 20 | Over 20 |
| --- | --- | --- | --- | --- | --- | --- |
| GI System | 5696 | 3630 | 285 | 44 | 28 | 22 |
| Cardiovascular System | 6126 | 3298 | 215 | 28 | 19 | 19 |
| Respiratory | 8713 | 960 | 32 |  |  | 0 |
| CNS | 7202 | 7202 | 2278 | 153 | 31 | 39 |
| Infections | 8856 | 846 | 2 | 1 |  | 0 |
| Endocrine | 2246 | 7307 | 143 | 7 |  | 2 |
| Obstetrics, Gynaecology and Urinary-tract | 8307 | 1386 | 9 | 3 |  | 0 |
| Nutrition and Blood | 8471 | 1185 | 31 | 14 | 2 | 2 |
| Musculoskeletal and Joint diseases | 9226 | 475 | 3 |  |  | 1 |
| Eye | 9425 | 278 | 2 |  |  | 0 |
| ENT | 9495 | 210 |  |  |  | 0 |
| Skin | 8016 | 1369 | 239 | 49 | 13 | 19 |
| Immunological products and vaccines | 9584 | 121 |  |  |  | 0 |
| Anaesthesia | 9687 | 18 |  |  |  | 0 |
| Malignant Disease and Immunosuppression | 9643 | 62 |  |  |  | 0 |

**Figure S1.** **Distribution of number of medications prescribed across different BNF chapters to individuals diagnosed with T1DM.**

Number of people with T2DM

| BNF chapters | 0 | 1 to 5 | 6 to 10 | 11 to 15 | 16 to 20 | Over 20 |
| --- | --- | --- | --- | --- | --- | --- |
| GI System | 48580 | 84401 | 9940 | 1503 | 781 | 702 |
| Cardiovascular System | 54476 | 81189 | 7945 | 1204 | 587 | 506 |
| Respiratory | 121277 | 23746 | 808 | 58 | 17 | 1 |
| CNS | 92802 | 48422 | 3217 | 759 | 339 | 368 |
| Infections | 130119 | 15670 | 112 | 6 |  | 0 |
| Endocrine | 51877 | 90135 | 3227 | 481 | 153 | 34 |
| Obstetrics, Gynaecology and Urinary-tract | 119542 | 26021 | 303 | 34 | 6 | 1 |
| Nutrition and Blood | 119430 | 25590 | 711 | 127 | 38 | 11 |
| Musculoskeletal and Joint diseases | 127996 | 17837 | 69 | 5 |  | 0 |
| Eye | 137458 | 8374 | 71 | 3 |  | 1 |
| ENT | 140836 | 5068 | 3 |  |  | 0 |
| Skin | 117977 | 22183 | 4125 | 1056 | 271 | 295 |
| Immunological products and vaccines | 143256 | 2650 | 1 |  |  | 0 |
| Anaesthesia | 145540 | 366 | 1 |  |  | 0 |
| Malignant Disease and Immunosuppression | 144197 | 1694 | 15 | 1 |  | 0 |

**Figure S2.** **Distribution of number of medications prescribed across different BNF chapters to individuals diagnosed with T2DM.**

**Table S1. The comorbidity group breakdown.**

| Morbidity categories | Co-morbidities |
| --- | --- |
| Alcohol substance abuse |  |
| Cancer |  |
| Cardiovascular/Cerebrovascular | CHD Atrial Fibrillation Heart Failure Hypertension Peripheral Vascular disease Stroke & Transient Ischaemic attack |
| Chronic Kidney disease |  |
| GI/ Liver disease | Chronic Liver disease Diverticular disease of intestine Inflammatory Bowel disease  Irritable bowel syndrome Constipation Dyspepsia Peptic Ulcer Disease |
| Glaucoma |  |
| Hypertension |  |
| Learning Disability |  |
| Mental Health Disorder | Anorexia bulimia Anxiety & other somatoform disorders Depression Schizophrenia or bipolar disorder |
| Dementia |  |
| Neurological Disorders | Migraine Epilepsy Multiple Sclerosis Parkinson's disease |
| Pain |  |
| Prostate Diseases |  |
| Respiratory/sinus | Asthma Bronchiectasis Chronic sinusitis COPD |
| Sensory | Blindness/Low vision Hearing Loss |
| Skin/Connective Tissue disorder | Psoriasis or Eczema Rheumatoid Arthritis & other inflammatory polyarthropathy/polyarthritis |
| Thyroid disorders |  |

**Table S2. Prevalence of comorbidity groups in individuals diagnosed with T1DM. The top 6 groups are shown in italics.**

| Morbidity group | N | % |
| --- | --- | --- |
| *Mental health disorder* | 4655 | 48.0 |
| *Hypertension* | 3767 | 38.8 |
| *GI/liver disease* | 3499 | 36.1 |
| *Pain* | 2493 | 25.7 |
| *Respiratory/sinus* | 2423 | 25.0 |
| *Cardiovascular/cerebrovascular* | 1562 | 16.1 |
| Sensory | 1554 | 16.0 |
| Skin/connective tissue disorder | 1378 | 14.2 |
| Neurological disorders | 1307 | 13.5 |
| Thyroid disorders | 1273 | 13.1 |
| Chronic kidney disease | 1035 | 10.7 |
| Alcohol/substance abuse | 773 | 8.0 |
| Glaucoma | 320 | 3.3 |
| Cancer | 242 | 2.5 |
| Prostate diseases | 245 | 2.5 |
| Dementia | 136 | 1.4 |
| Learning disability | 107 | 1.1 |
